# Faltering mortality improvements at young-middle ages in high-income English-speaking countries

**DOI:** 10.1101/2023.11.09.23298317

**Authors:** Sergey Timonin, David A. Leon, Emily Banks, Tim Adair, Vladimir Canudas-Romo

**Author notes:** **Corresponding Author:** Dr Sergey Timonin, School of Demography, College of Arts and Social Sciences, The Australian National University, 2601 Acton, Canberra, ACT, Australia.

## Abstract

**Background:** Before the COVID-19 pandemic, stagnating life expectancy trends were reported in some high-income countries (HICs). Despite previous evidence from country-specific studies, there is a lack of comparative research that provides a broader perspective and challenges existing assumptions. This study aims to examine longevity trends and patterns in six English-speaking countries (Australia, Canada, Ireland, New Zealand, UK, USA) by combining period and cohort perspectives and to compare them with other HICs.

**Methods:** Using data from the Human Mortality and WHO Mortality Databases, we estimated partial life expectancy, lifespan inequality and cohort survival differences for 1970-2021, as well as the contribution of causes of death to the gap in life expectancy between English-speaking countries and the average for other HICs in 2017-19.

**Results:** In the pre-pandemic period, the increase in life expectancy slowed in all English-speaking countries, except Ireland, mainly due to stagnating or rising mortality at young-middle ages. Relative to other HICs, those born in Anglophone countries since the 1970s experienced relative survival disadvantage, largely attributable to injuries (mainly suicides) and substance-related mortality (mainly poisonings). In contrast, older cohorts enjoyed advantages for females in Australia and Canada and for males in all English-speaking countries except the USA.

**Conclusions:** Although future gains in life expectancy in wealthy societies will increasingly depend on reducing mortality at older ages, adverse health trends at younger ages are a cause for concern. This emerging and avoidable threat to health equity in English-speaking countries should be the focus of further research and policy action.

**Key messages:**

- The study highlights striking similarities between English-speaking high-income countries in terms of adverse health outcomes at young-middle ages compared to a group of other high-income countries (HICs).
- Each of the Anglophone populations has experienced a marked mortality disadvantage for cohorts born since the early 1970s relative to the average of other HICs, which contrasts with the generally better performance of the older cohorts in some English-speaking countries, particularly for men.
- In the most recent pre-pandemic period, i.e. 2017-19, the negative contribution of higher mortality at ages below 50 years to the gap in life expectancy at birth between English-speaking countries (excluding Ireland) and other HICs ranged from 0.15 years for Australian women to 2.06 years for US men.

## Background

Global life expectancy has grown dramatically over the past century, although the underlying declines in mortality have not been uniform between and within populations.^1,2^ In the years preceding the COVID-19 pandemic, stagnations or even reversals in longevity trends were reported in some high-income countries (HICs),^3–8^ which are warning signs of both long-standing and emerging health problems.^9^ The COVID-19 pandemic has had a further impact on global mortality, including in the HICs, adding a degree of uncertainty to future trends in life expectancy.^10,11^

Much of the recent discourse on life expectancy trajectories has shifted towards the growing challenge of mortality in old age in rapidly ageing societies. A comparative study showed a substantial, albeit temporary, decline in life expectancy in most HICs in 2014-15, which was likely related to a particularly severe influenza season, and was predominantly driven by changes in mortality at older ages.^12^ The subsequent COVID-19 pandemic has had the largest mortality impact on older populations in HICs, with the exception of Eastern Europe and the USA, where working age adults also experienced a severe increase in mortality.^11,13^ In addition to the direct and indirect effects of seasonal viruses, austerity policies, weaknesses of health and social care systems, and growing socio-economic disparities within countries are seen as key challenges to further sustained improvements in longevity.^7,14–19^

The stagnation of life expectancy in the USA and the UK, the two worst affected high-income countries, is largely, though not exclusively, related to negative mortality trends in midlife.^20–22^ Although the causes of the adverse health trends in these two English-speaking (or Anglophone) countries differ, one of the similarities is the growing burden of so-called “deaths of despair” (refer to drug- and alcohol-related deaths plus suicides) which was first thought to be a US phenomenon^23^ but has since been observed in the UK as well.^16^ Examining substance-related mortality in the USA, Ho^24^ found disturbing patterns of drug overdose in other English-speaking countries (namely, Australia and Canada), which was further explored by Dowd et al.^25,26^ The opioid death crisis in Canada has been highlighted elsewhere.^27^ It has also been shown that the USA and Australia have some of the highest adult obesity prevalence rates among HICs, which was suggested to be a key factor in the recent stalling of CVD mortality in both populations.^28,29^

These observations have led us to hypothesise that English-speaking high-income countries may share some common patterns in recent mortality changes and some fundamental similarities (as well as differences) in long-term trends. Furthermore, contrasts with other HICs may yield clues for improvement. In this study, we aim to conduct a comparative study that examines longevity trends and patterns in six English-speaking countries in relation to an average of 14 other HICs. First, we explore the long-term time trends in life expectancy and lifespan inequality. We then assess the contribution of ages and causes of death to life expectancy gap in the recent pre-pandemic period. Finally, the differentials in cohort survival between English-speaking and peer countries are examined.

## Methods

Our analysis focuses on six English-speaking high-income countries (Australia, Canada, Ireland, New Zealand, the UK, and the USA) in comparison with an average of 14 other HICs. The comparator group includes high-income, low-mortality countries from Western Europe plus Japan (Supplementary Table S1), following a similar approach to others.^6,21,30^ The additional (sensitivity) analysis uses a different comparison group of five best-performing HICs (Japan, Switzerland, Italy, Spain and Sweden).

We used the data from well-established, comprehensive sources of high-quality mortality data: Human Mortality Database (HMD)^31,32^ for all-cause mortality data for 1970-2021, and the WHO Mortality Database (WHO MB)^33^ for cause-specific death counts for 2017-19.

We estimated annual life expectancy at birth, partial life expectancy between ages 0 and 50 years, remaining life expectancy at age 50 and lifespan disparity for each English-speaking country and the comparator group. While life expectancy is an average mortality measure, lifespan disparity measures variation in age-at-death and helps to capture trends associated with deteriorating mortality, particularly at younger ages.^14,34–36^ To assess the temporal dynamics of life expectancy at birth, we calculated its mean annual changes for historical periods (1970-90, 1990-2010), the pre-pandemic decade (2010-15, 2015-19) and the pandemic period (2019-21). We further decomposed^37^ these changes into the age-specific contributions.

To explore the cohort survival differences between Anglophone and other HICs, we calculated and plotted the gap in the truncated cross-sectional average length of life (TCAL).^38^ From a public health perspective, this approach allowed identifying birth cohorts exposed to higher/lower historical mortality.^30^

To assess the impact of causes of death on the life expectancy gap between Anglophone populations and other HICs, we applied decomposition techniques^37,39^ to quantify the contribution of ages and causes to the observed differences in 2017-19. The three-year average was used to avoid random fluctuations and to ensure the robustness of our estimates. Recognising the variation in death certification practices between countries, we categorised deaths by assigning ICD-10 codes to create cause groups of public health and clinical relevance, in line with previous analyses (Supplementary Table S2).

All analyses were performed in R Studio. More detailed information on data handling and methods can be found in the Supplementary Data and Methods.

## Results

### Long-term trends in life expectancy and lifespan disparity

Figure 1 shows the relationship between trends in average longevity, as measured by life expectancy at birth, and lifespan disparity, which reflects the variation in age at death. In addition to the previously documented rise in lifespan inequality in the USA^40^, there have also been increases in lifespan variation in other English-speaking countries, suggesting growing health problems at younger ages in these populations. In the comparator group, the gains in life expectancy coincided with a further decline in lifespan variation.

**Figure 1.**
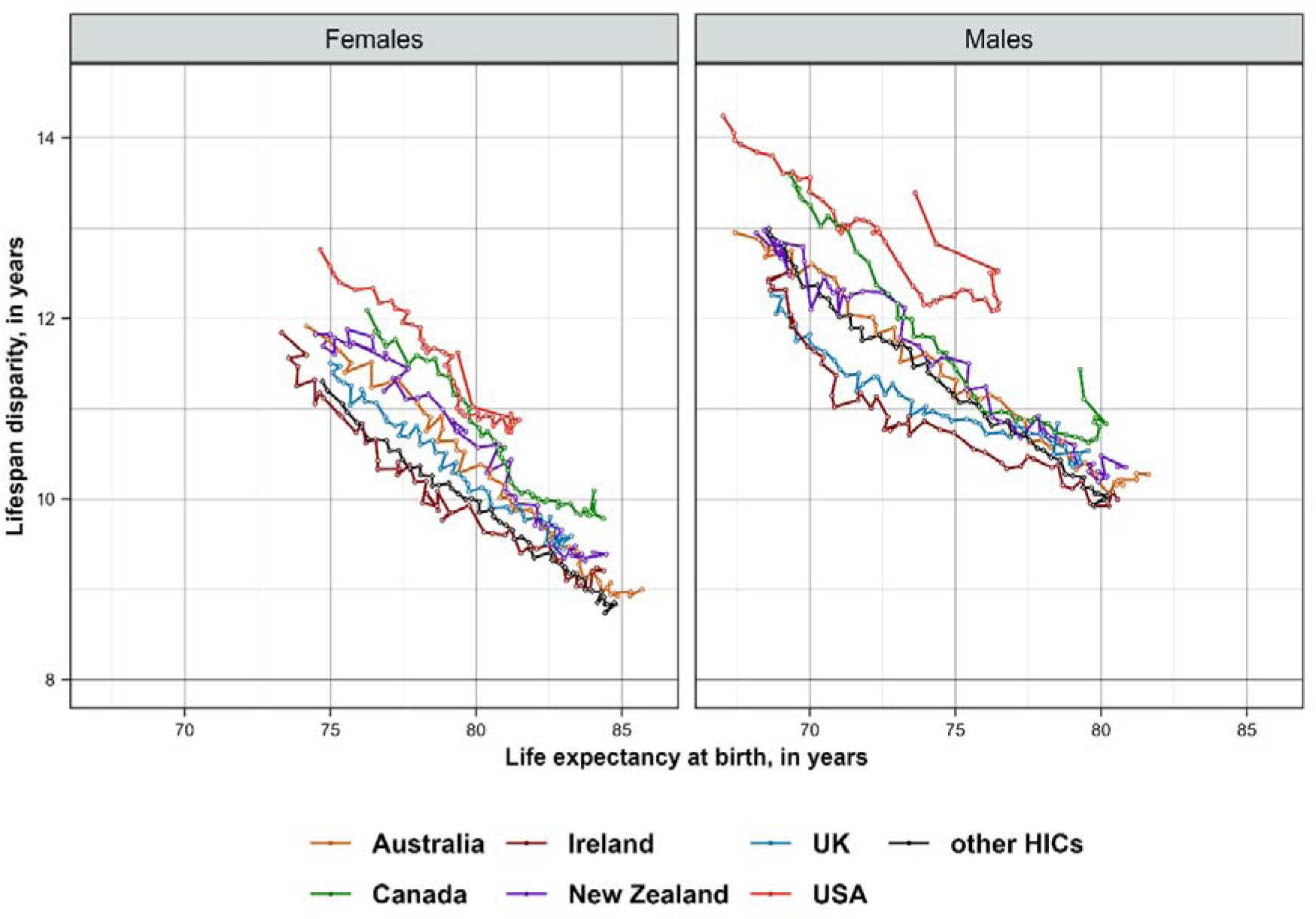
Associations between changes in life expectancy at birth and lifespan disparity in English-speaking countries and the average for other HICs, by sex, 1970–2021. Notes: ^a^ HMD, used as the primary data source for the calculations, does not yet contain data for 2021 for four countries (see Supplementary Table S1 for more information on data availability).

Figure 2 separates the sex-specific trends in life expectancy at birth into trends in partial life expectancies between ages 0 and 50 years (upper panel), and at age 50 years (lower panel). In terms of remaining life expectancy at age 50, men in all English-speaking countries (except for the USA) perform better or similar to the average of other HICs, while this only occurred for women in Australia and Canada. In contrast, the life expectancy at young-middle ages of Anglophone countries began to lag behind the reference group in the mid-2000s. Supplementary Figure S1, similar to Figure 2 but using the average of the five best-performing other HICs, further highlights the life expectancy disadvantage of all English-speaking countries at ages below 50 and the relative advantage of Australian males at older ages, which puts them close to the world record.

**Figure 2.**
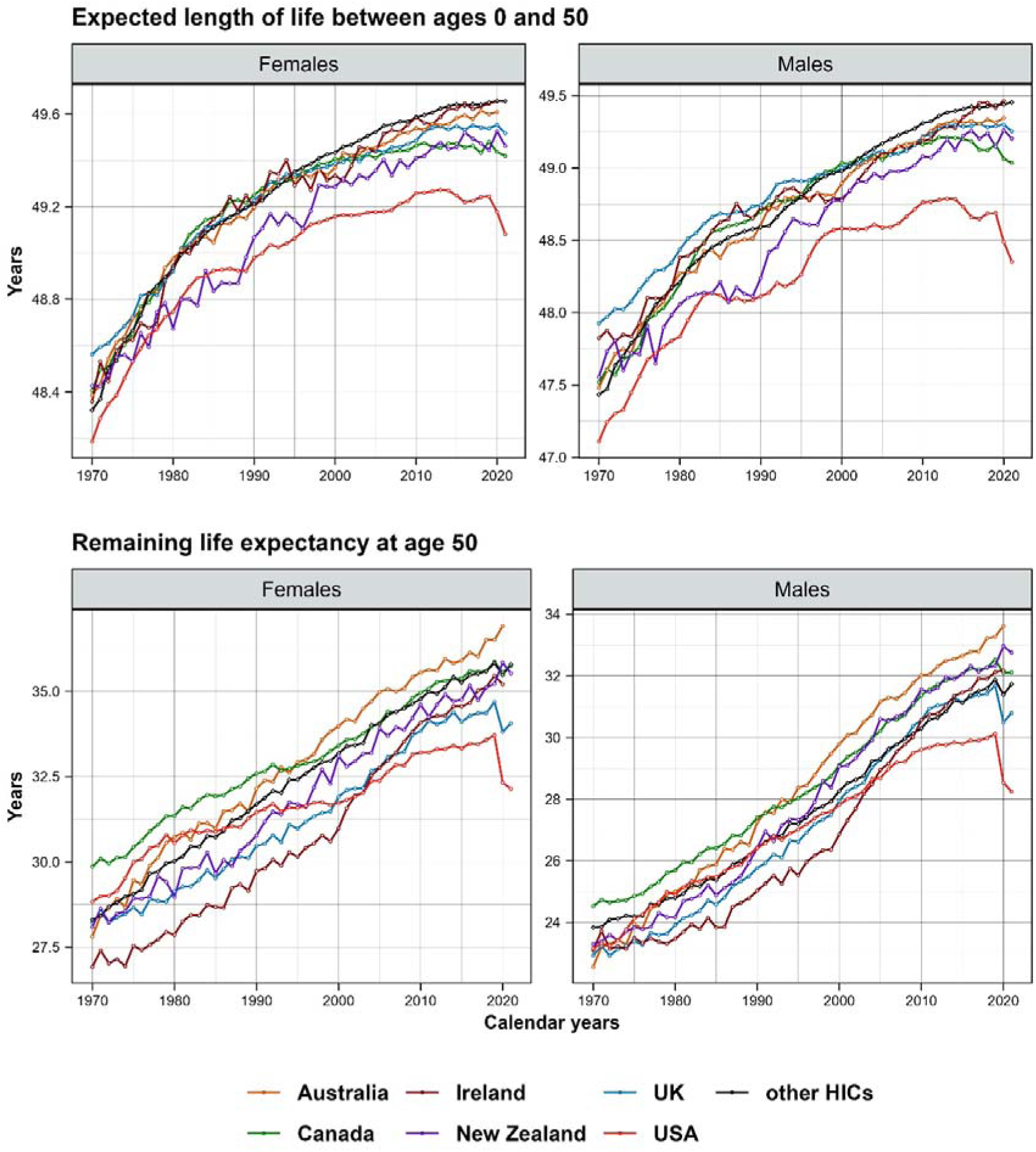
Time trends in partial life expectancies in English-speaking countries and the average for other HICs, by sex, 1970-2021. Notes: ^a^ life expectancy at birth is the sum of life expectancies between ages 0 and 50 (upper panel), and remaining life expectancy at age 50 (lower panel) multiplied by the probability of surviving up to age 50 (see Supplementary Methods); b HMD, used as the primary data source for the calculations, does not contain data for 2021 for four countries (see Supplementary Table S1 for more information on data availability).

Supplementary Figure S2, which shows the association between life expectancy below and above age 50 for each HIC separately, further confirms the relative disadvantage of the English-speaking countries (except for Ireland) in mortality at young-middle ages.

### Recent stagnation in longevity improvements across English-speaking countries and diverging trends during the COVID-19 pandemic

Figure 3 compares the contribution of three age groups (0-4, 5-49, and 50+ years) to the mean annual changes in life expectancy at birth in recent periods (2010-15, 2015-19, and 2019-21) with the earlier periods of 1970-90 and 1990-2010. For each of the Anglophone countries, the smallest gains in life expectancy were recorded in the period between 2010 and 2015 (except for Irish males). They were also smaller than the average for the other HICs. In the next period, 2015-19, gains in life expectancy were higher for all female populations, but remained unchanged or were even smaller for males. Only females in Australia and Ireland had a higher increase in life expectancy than the reference group.

**Figure 3.**
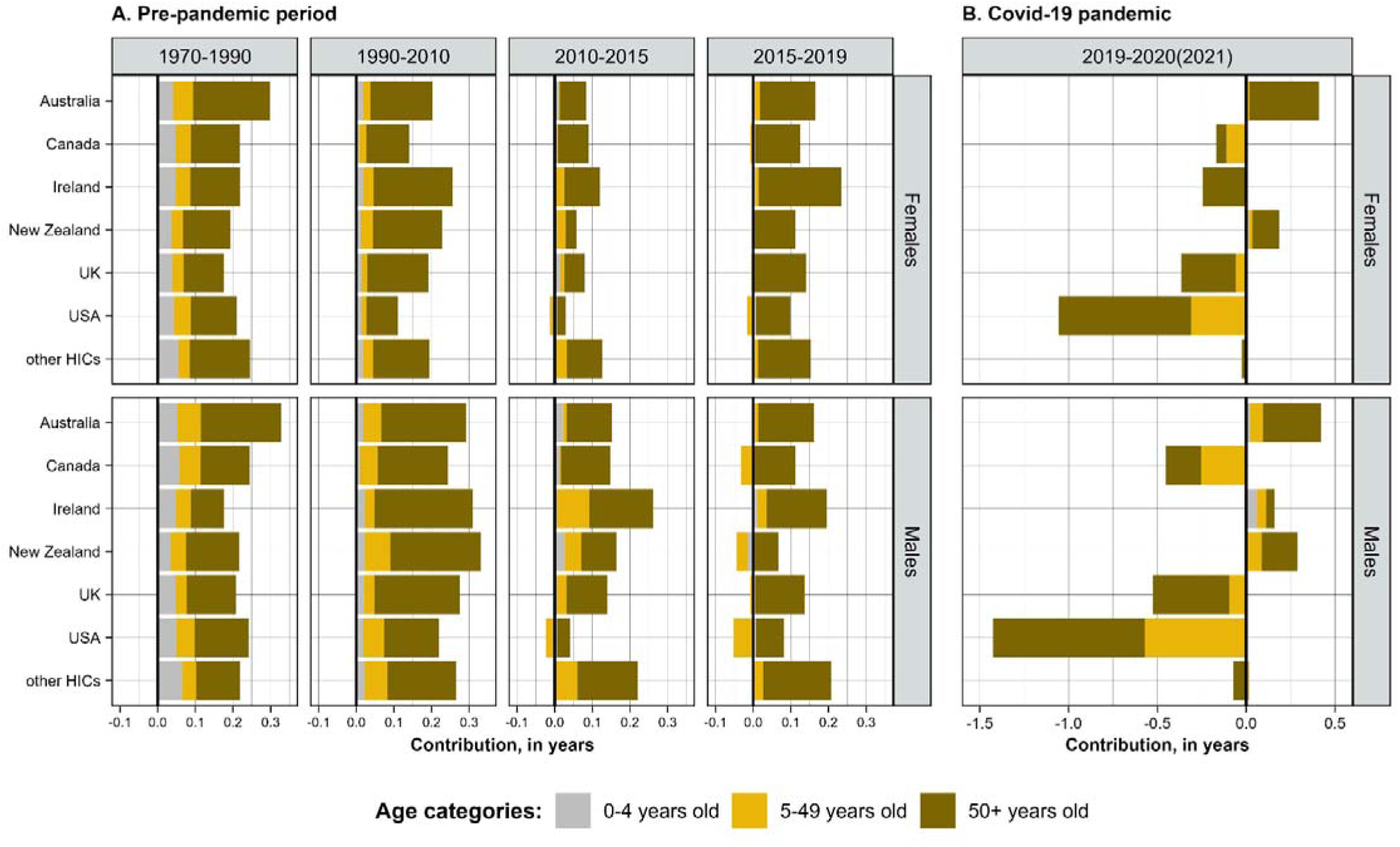
Age-specific contributions to the mean annual changes in life expectancy at birth in three pre-pandemic periods (1970-2010, 2010-15, and 2015-19) and during the COVID-19 pandemic (2019-21), by sex, in years. Notes: Positive values correspond to age groups contributing to life expectancy increase, and negative values contributing to its decrease; the corresponding values are presented in Supplementary Table S3. Input data for Australia and Ireland are not available for 2021.

The contribution of mortality changes at young-middle ages has generally declined over time in all populations, while the role of older ages has become more pronounced. Importantly, mortality changes between ages 5 and 50 had a negative impact on life expectancy trends for both sexes in the USA in 2010-15 and 2015-19, for males in Canada and New Zealand and both sexes in the UK in 2015-19.

The COVID-19 pandemic had a differential effect on life expectancy change in English-speaking countries (Figure 3). It has had the largest negative impact in the USA, the UK, and Canada, including their young-middle-aged groups. The average decrease in life expectancy in a group of other HICs was smaller and mainly affected older people.

### The contribution of ages and causes of death to the life expectancy gap

The results of the age- and cause-specific decomposition of the gap in life expectancy at birth between English-speaking countries and the average for other HICs are shown in Figure 4 and Supplementary Table S3. In 2017-19, males and females in Australia had an overall longevity advantage over the comparator group, as did the male populations of Ireland, New Zealand and Canada. The rest of the English-speaking populations had lower life expectancy at birth compared to the average for other HICs.

**Figure 4.**
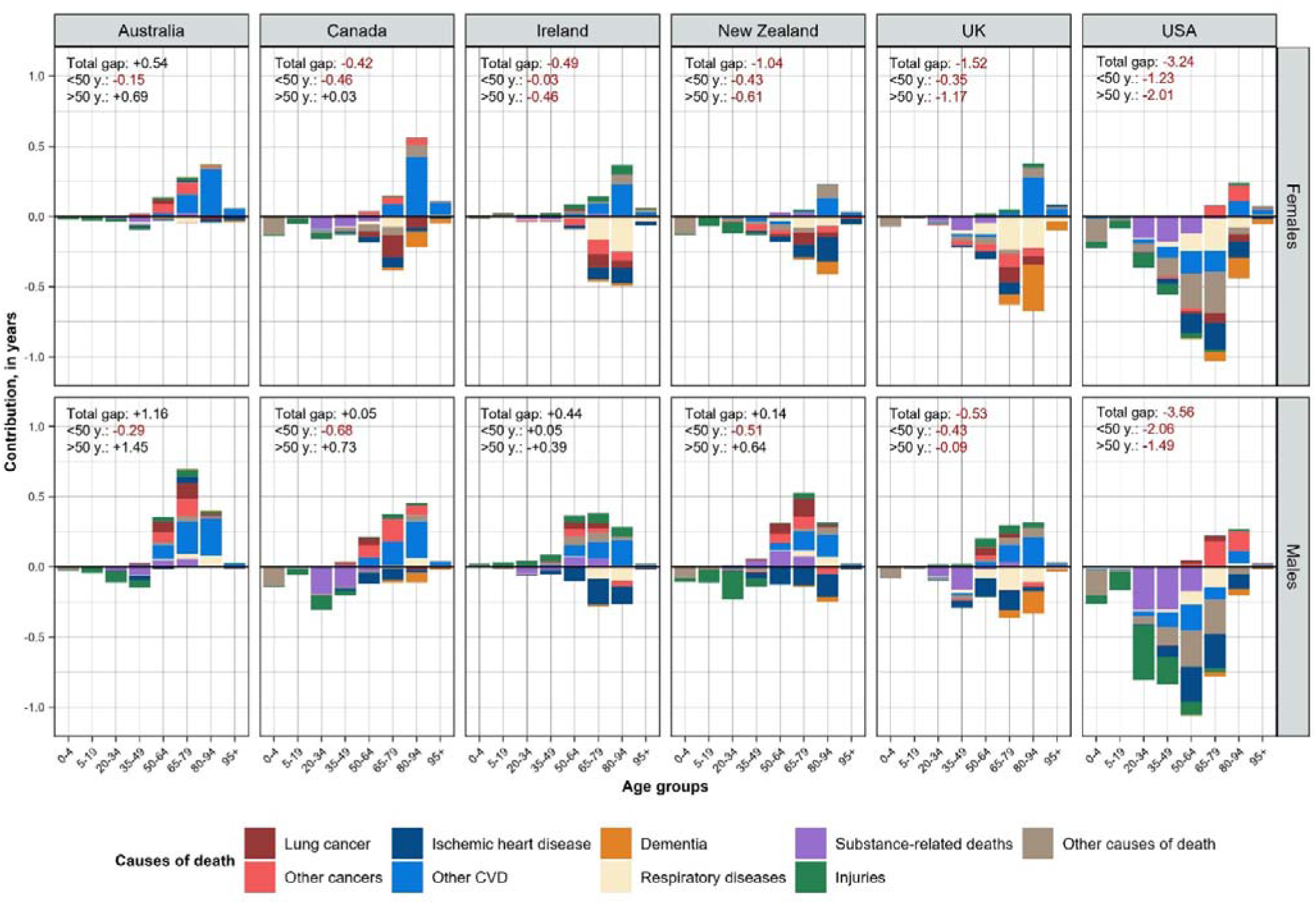
Age- and cause-specific contributions to the gap in life expectancy at birth between each of the English-speaking countries and the average for other HICs, 2017-19, by sex. Note: positive values correspond to ages and causes that contribute to higher life expectancy in English-speaking countries compared to other HICs; negative values contribute to lower life expectancy than the other HICs. The corresponding values (for each cause of death and broad age intervals) are presented in Supplementary Table S3.

However, the striking similarity across all English-speaking countries was that mortality before age 50 made a negative contribution to the gap in life expectancy at birth between each Anglophone country and the comparator group (except for Irish men, where this contribution was close to zero). The contribution of mortality at older ages did not show a clear tendency across countries, with one nuance - older men in English-speaking countries were more likely than women to have lower mortality than the average for other HICs.

The contribution of causes of death to the differences in life expectancy was quite diverse across ages and countries (Figure 4). Injuries and substance-related deaths (mainly suicides and poisonings) accounted for most of the losses of life expectancy in English-speaking countries at young-middle ages. For older males, lower mortality from both cardiovascular diseases and cancer were the two main contributors to the life expectancy advantage of some English-speaking countries. For females, only CVD mortality had a clear positive impact on life expectancy advantage in Australia and Canada, while higher mortality from cancer, dementia, and respiratory diseases (incl. COPD) increased the longevity disadvantage for other Anglophone countries.

### Differential cohort survival

Differences between the English-speaking countries and the comparator group in the cumulative survival of birth cohorts are shown in Figure 5. Compared with the reference group, there is a clear survival advantage for men born in the 1930-40s in Australia, Canada and New Zealand, and for those born around the 1950s in Ireland and the UK. For women, however, a strong cohort survival disadvantage is observed in Ireland, New Zealand, the UK, and Canada (1930s birth cohorts), with only Australian women outperforming the average of other HICs. In the USA, there has been a growing disadvantage for all cohorts in both sexes, except for the very old populations born in the 1930s.

**Figure 5.**
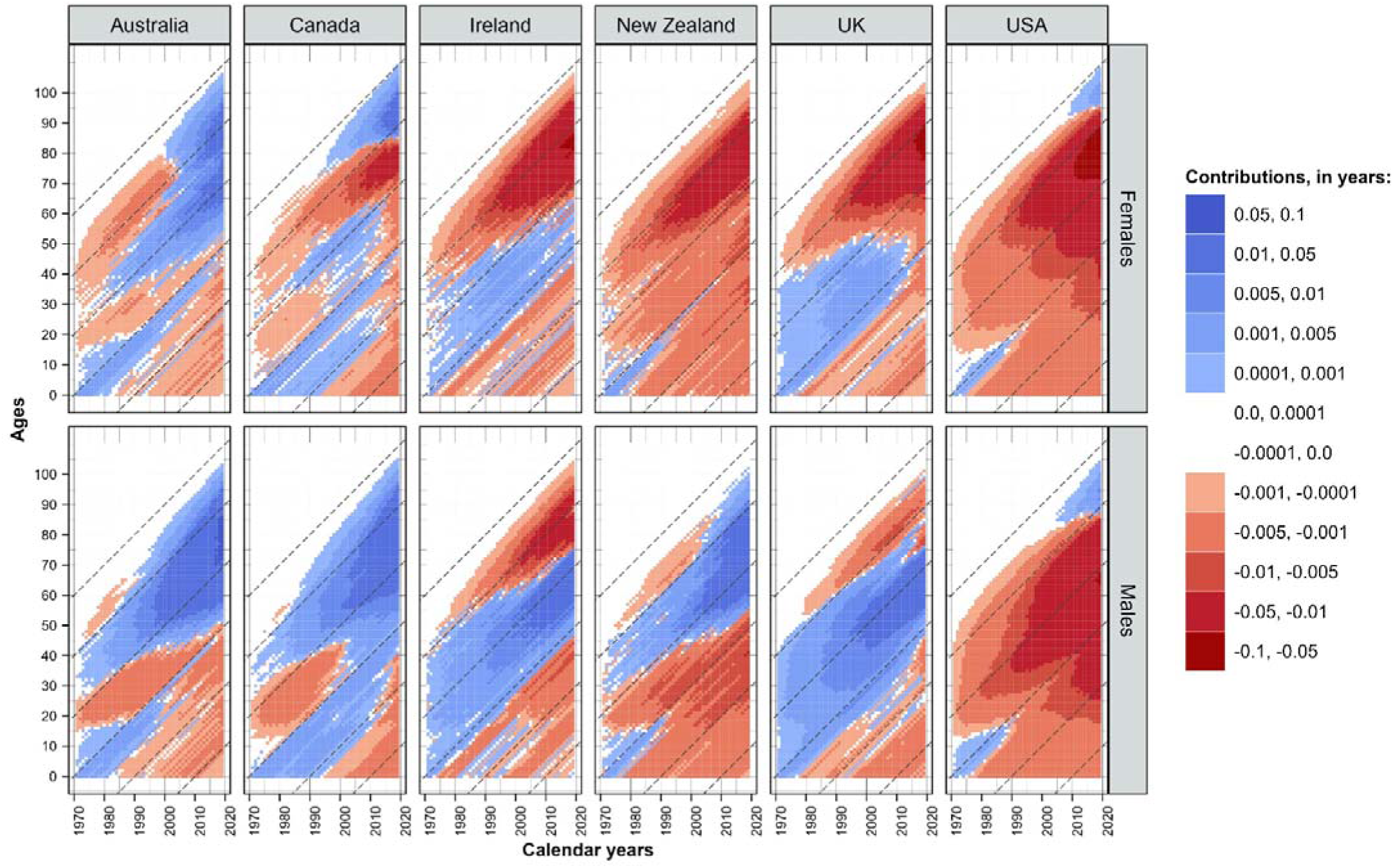
Cumululative differences in cohort survival between each of the English-speaking countries and the average for other HICs, by sex, from 1970 to 2019. Notes: positive values correspond to higher cohort survival in English-speaking countries while negative values favour the other HICs. The black diagonal lines indicate different birth cohorts born in 1910, 1930, 1950, 1970, 1990, and 2010.

The red triangles in the lower right-hand corner of each plot in Figure 5 reflect the accumulated mortality disadvantage of birth cohorts born in the early 1970s and later. In other words, these male cohorts in English-speaking countries have experienced higher mortality than the comparison group, with a similar but less pronounced pattern for females. These results reinforce the findings in the previous section of the negative contribution of injuries and drug-related deaths among young and middle-aged adults to the gap in life expectancy at birth in the recent pre-pandemic period.

## Discussion

Our analysis has uncovered several important aspects of mortality patterns and trends in a group of English-speaking countries. First, in the decade before the pandemic, most HICs experienced a slowdown in life expectancy gains, which was particularly marked in English-speaking countries. Anglophone populations (except for Ireland) had a smaller increase in life expectancy in 2010-19 compared with the average for other HICs and with the trends observed in previous decades. This was largely the result of lessening improvements or even significant rises (in the USA in 2010-19, and in Canada, the UK and New Zealand (males only) in 2015-19) in mortality at ages below 50. Second, these adverse mortality trends in Anglophone countries resulted in negative contributions of the under-50 age group to the overall life expectancy gap with the comparator group in 2017-19. The observed health disadvantage of English-speaking countries at this age was mainly due to mortality from injuries (including suicides), and drug and alcohol poisonings. Third, cohort analysis has shown that males born in English-speaking countries since the 1970s have had lower survival rates than the average of other HICs. In contrast, older male cohorts have enjoyed lower mortality, except for the USA. For females, with the exception of Australia and Canada at older ages, survival has been lower in English-speaking countries than in other HICs over the whole range of ages and birth cohorts.

To our knowledge, this is the first comparative study examining longevity trends in a group of English-speaking high-income countries from both a period and a cohort perspective. The relative mortality disadvantage of each Anglophone country (expect for Irish males) at young-middle ages is the most striking and important observation. Another common feature is that, compared to the average of the other high-income countries, the gap in life expectancy at birth for males in Anglophone countries was smaller than females. In contrast, the negative contribution of mortality patterns at ages below 50 years to the overall life expectancy gap was larger for males than for females. External causes of death and substance use disorders were found to be the largest contributors to the loss of life expectancy between the ages of 5 and 50 years in all English-speaking countries. Higher mortality from CVD (particularly in males) as well as from neoplasms and respiratory diseases in the female populations of New Zealand and the UK also played an important role at these ages.

In general, the results of our study are consistent with some of the previous findings from the range of country studies. They are also in a line with earlier observations of slowing mortality improvements in a larger number of high-income/OECD countries,^4,6^ and with negative trends in drug overdose mortality in some of the English-speaking countries.^24,25,27^ However, our study went further by dissecting a group of English-speaking countries from various perspectives, rather than just looking at individual populations or the whole range of high-income countries. Even if the USA still appears to be an anomaly among rich countries (in terms of adverse mortality trends), other high-income countries, and especially Anglophone countries, are not guaranteed to be immune to the factors underlying “American disease”.^41^

The fact that other HICs have not experienced the same adverse mortality trends in young-middle-aged adults should provide insights into larger-scale drivers of health and potential improvement. While the exact reasons for differences cannot be determined from the data presented, promising candidates include measures that reduce inequity and structural disadvantage, that address obesity, firearms and violence, and that support mental health and drug-related safety.

In addition to using the most reliable, carefully harmonised and validated mortality data, the main strength of our study was the use of both period and cohort methods to provide a comprehensive analysis of both recent and long-term trends in mortality. We used traditional and widely accepted measures of longevity such as life expectancy at different ages as well as more novel metrics, such as lifespan inequality, and truncated cross-sectional length of life, to address the research question.

## Limitations

Our study has some limitations. The comparator group of other HICs is not homogeneous. However, Supplementary Figure S2 shows that the main conclusions based on the comparison with the average for other HICs are generally consistent when analysing individual countries. The additional decomposition analysis, using the five-best performing HICs as a comparison group, has also highlighted the striking mortality disadvantage of English-speaking countries at young-middle ages (Supplementary Figure S3).

Although our study included countries with high-quality mortality statistics, the cause-of-death comparisons may be subject to some inconsistencies related to differences in coding practices across countries.^42,43^ Rather than analysing time trends in cause-specific mortality, we focused only on assessing the contribution of causes of death to the gap in life expectancy between Anglophone and other HICs in the most recent pre-pandemic period. However, it may be important in the future to examine the contribution of causes of death to the cohort survival differences between the countries.^44^ The use of TCAL as a summary measure of the cohort survival experience of a population has the advantage of using all the available mortality data (1970-2021) for each population, avoiding any possible cohort identification problem raised in a recently published paper.^45^

We deliberately did not focus much on trends during the COVID-19 pandemic for two main reasons. First, it would add a layer of complexity and obscure our main findings. Second, final data for 2020-22 are not yet available for some countries. However, as briefly shown in the results section and elsewhere,^11,46,47^ the impact of the pandemic on the HICs varied considerably in terms of age (stronger negative effect on young-middle-aged mortality in the US, UK and Canada) and timing (shift of the peak of excess mortality to 2021 and even 2022 in Australia and New Zealand). Further analyses should be carried out when the final data are published.

While we did not aim to provide a comprehensive analysis of the potential factors that might explain the observed slowdown in survival improvements, this paper examined a previously overlooked mortality disadvantage of all high-income English-speaking countries at young-middle ages through the lens of period and cohort analysis.

## Conclusion

Although future gains in life expectancy in high longevity societies will increasingly depend on reducing mortality in old age, patterns of mortality in young and middle-aged adults are a cause for concern. The persistence or increase in mortality at these ages may also be an important indicator of other non-fatal health problems, including mental disorders, and requires further investigation. Detailed causes-specific analysis (including multiple causes of death) and examination of socioeconomic inequalities in health in the English-speaking countries are likely to shed further light on some of these issues. There is scope for the English-speaking countries to improve the health of their younger populations and to halt the widening gap in mortality with other high-income countries.

## Ethics approval

Ethics approval was not required because the study is an analysis of aggregate data.

## Data availability

The raw data originated from the publicly available sources listed in the references. The analytical codes used to perform the statistical analysis, as well as instructions on how to access the data, can be obtained on reasonable request to the corresponding author at.

## Supplementary data

Supplementary data are available at IJE online

## Author Contributions

S.T. and V.C-R. conceived the study. S.T. collected, pre-processed and validated the data. S.T. carried out the statistical analyses with support from V.C-R. S.T. wrote the first draft. All the authors provided critical feedback on the manuscript, contributed to subsequent versions, and to the interpretation of the data and results. All the authors reviewed and approved the final version of the manuscript.

