## Supplementary Materials for "Faltering mortality improvements at young-middle ages in high-income English-speaking countries"

**Supplementary Data and Methods**

### Supplementary Data. Data sources and data handling

For the all-cause analysis, we relied on 1-year age- and sex-specific death rates with the open-end age interval 110 years and above. These data were extracted from the Human Mortality Database (HMD) for each country (both English-speaking and the peers) for 1970-2021.^1^ We used the data on Eastern and Western Germany separately for the period from 1970 to 1989 because it was not available for the whole territory of Germany.

Age- and sex-specific death rates for the comparator group, denoted with a bar on top of the variable, were estimated as the unweighted average of country-specific rates

$\bar{m}_{x,s}=\frac{\sum_{i=1}^{k} m_{x}^{i}}{k}$,

where $m_{x,s}^{i}$ is the central (all-cause) death rate for age *x,* sex *s* and country *i;* and *k=14* the total number of countries in the comparator group.

We used mean values instead of, for instance, median estimations because this approach was considered as the most suitable and transparent particularly when cause-specific analysis was conducted. Moreover, our sensitivity analysis (not shown here) confirmed minor differences between mean and median estimations of longevity measures.

For cause-specific analysis we extracted death counts from the WHO Mortality Database for the most recent pre-pandemic period, i.e. 2017-2019.^2^ A three-year average was used to obtain more robust estimations and to avoid potential short-term fluctuations. However, cause-specific death counts were not available for Norway for the whole period under consideration (Supplementary Table S1). In these cases, our estimates of cause-specific proportions were based on data for available years. The cause-of-death data for New Zealand (2017-19) was sourced from the Health New Zealand Mortality Collection.^3^

The cause-specific data required some addition preparation before it could be used for the purposes of statistical analysis. For those country-years with the last age interval 85+ years (Canada, Ireland, Finland, France, Germany, and New Zealand), we apply the ‘ungroup’ R package to get the age distribution up to age 95+.^4^

We then estimated the average for 2017-19 cause-specific proportions of total deaths for each age, sex and country as follows:

$R_{x,s}^{i, j}=\frac{D_{x,s}^{i, j}}{D_{x,s}^{i}}$,

where $D_{x,s}^{i, j}$ is the death counts for age *x,* sex *s* and country *i,* and cause of death *j*.

Finally, we obtained the cause-specific death rates by multiplying the proportions by corresponding central death rates (0, 1-4, 5-9, …, 95+) estimated from the HMD as

$m_{x,s}^{i, j}= R_{x,s}^{i, j}*m_{x,s}^{i}$.

We made some additional adjustments to obtain the average cause-specific death rates (and proportions) for the comparator group. The estimated cause- and age-specific death rate for the comparator group is simply the mean of the cause-, age- and country-specific death rates:

$${\hat{\bar{m}}}_{x,s}^{j}= \frac{\sum_{i=1}^{k} m_{x,s}^{i, j}}{k}.$$

The final value is adjusted by the age-specific coefficient as

$\bar{m}_{x,s}^{j}={\hat{\bar{m}}}_{x,s}^{j}* \frac{\bar{m}_{x,s}}{\sum_{j=1}^{r} {\hat{\bar{m}}}_{x,s}^{j}}$,

where *r* is the number of causes of death.

### Supplementary Methods. Statistical and demographic analysis

*Life table construction.* We used standard methods^5^ to construct complete period life tables using the 1-year (up to 110+) age-, sex-, and country-specific death rates. From the life tables, we extracted life expectancy at birth ($e_{0}$), expected length of life between ages 0 and 50 (${{}_{50}e}_{0}$), and the remaining life expectancy at age 50 ($e_{50}$). In continuous notation, these life table functions can be expressed as:

$e_{0}=\int_{0}^{110} l\left( x \right)dx; {{}_{50}e}_{0}=\int_{0}^{50} l\left( x \right)dx;e_{50}=\frac{\int_{50}^{110} l\left( x \right)dx}{l(50)}$,

where *l(x)* is the life table survival function (number alive at age *x* from an initial synthetic cohort); and the radix of the life table is *l(0)* = 1.

Following the formulas above, life expectancy at birth, between ages 0 and 50, and remaining life expectancies are related in the following way:

$e_{0}={{}_{50}e}_{0}+l\left( 50 \right) e_{50}$.

The age of 50 was chosen as the threshold age after a careful preliminary analysis of age-specific mortality trends in English-speaking countries compared to other HICs.

In addition to using average longevity measures such as life expectancy, we also estimated the lifespan disparity metric (e-dagger or e†). It shows the variation (dispersion) in age at death and can also be interpreted as the average years of life lost due to premature mortality. This measure is particularly useful (to complement to life expectancy indices) to monitor the emergence of health crisis, particularly among younger populations (premature deaths). The continuous^6^ and discrete^7^ formulas for lifespan disparity ($e^{\dagger}$) from a complete period life table is as follows:

$e^{\dagger} = \int_{0}^{110} e\left( x \right)d\left( x \right)dx\boldsymbol{\cong} \sum_{x=0}^{109} [d_{x}*\left( e_{x}+a_{x}*\left( e_{x+1}-e_{x} \right) \right]+d_{110+}*e_{110+}$,

where$d\left( x \right)$ is the life table distribution of deaths; $d_{x}$ and $a_{x}$ are the life table number of deaths and share of the age interval lived by those dying in the age interval from *x* to *x+1*.

*Age- and cause-specific decomposition of life expectancy differences/changes.* Decomposition techniques allow differences in life expectancy between two populations, or its change between two points in time, to be broken down into the contribution of age and causes of death. To perform this task, we followed the stepwise replacement approach.^8^ This algorithm estimates the effect of replacing each elementary cell in one matrix (age- and/or cause-specific death rates in population 1) with the corresponding cell in the second matrix (age- and/or cause-specific death rates in population 2). We used the package ‘DemoDecomp’ to conduct decomposition analysis.^9^

*Truncated cross-sectional average length of life (TCA*L). We calculated the TCAL(t) measure to estimate cohort survival differences between English-speaking countries and other HICs. TCAL is similar to the period life expectancy at birth with one important difference - it takes into account not only the age-specific mortality pattern in a given year, but also the survival history of the cohorts present in a calendar year for which this measure is estimated.^10^ In other words, it is a period life expectancy measure adjusted for the historical mortality of the cohorts from their birth or the age at which the data first became available.

We computed TCAL for the last pre-pandemic year (i.e. 2019) using the data starting in 1970 to capture the mortality patterns over this period:

$TCAL (t, Y)=\int_{0}^{110} l\left( x, t,Y \right)dx$,

where *t* = 2019, *Y* = 1970, *l(x,t,Y)* - the survival function for cohorts born in year *t-x* reaching age *x* in 2019.

The survival function was calculated as the product of single age probabilities to survive from age *0* to *x.* For cohorts born before 1970 we assumed death rates equal to zero focusing our comparisons only on the period since 1970. The year of truncation corresponded to the overall time period of our analysis in this study.

The differences in TCALs between each of the English-speaking countries and the comparator group were further decomposed into age- and cohort-specific contribution as:

$\Delta(x, c,{ENG}_{i})=\left[ \frac{l\left( x,t,Y,{ENG}_{i} \right)+l\left( x,t,Y,HIC \right)}{2} \right] ln\left[ \frac{{}_{1}{p_{x}}\left( c,{ENG}_{i} \right)}{{}_{1}{p_{x}}\left( c,HIC \right)} \right]$,

where *c* – birth cohort (i.e. *t*-*x*), *ENG_i_* – each of the English-speaking countries, HIC – high-income countries (comparator group), *_1_p_x_(c)* – probability of surviving from age *x* to *x+1* for the cohort *c* (i.e. born in year *t-x*).

The age-specific contributions in year *t* (i.e. 2019 in our case) is the cumulative sum of contributions across each cohort reaching age *x* in year *t*.

### Table S1. Study populations and data availability by data source (as of 01.06.2024)

|  | ***Country name*** | ***Country group*** | ***HMD data availability*** | ***WHO data availability*** |
| --- | --- | --- | --- | --- |
| 1 | Australia | English-speaking countries | 1970-2020 | 2017-2019 |
| 2 | Canada |  | 1970-2021 | 2017-2019 |
| 3 | Ireland |  | 1970-2020 | 2017-2019 |
| 4 | New Zealand |  | 1970-2021 | 2017-2019 ^a^ |
| 5 | United Kingdom |  | 1970-2021 | 2017-2019 |
| 6 | United States of America |  | 1970-2021 | 2017-2019 |
| 7 | Austria | Comparator group | 1970-2019 | 2017-2019 |
| 8 | Belgium |  | 1970-2021 | 2017-2019 |
| 9 | Denmark |  | 1970-2021 | 2017-2019 |
| 10 | Finland |  | 1970-2021 | 2017-2019 |
| 11 | France |  | 1970-2021 | 2017-2019 |
| 12 | Germany ^b^ |  | 1970-2020 | 2017-2019 |
| 13 | Italy * |  | 1970-2021 | 2017-2019 |
| 14 | Japan * |  | 1970-2021 | 2017-2019 |
| 15 | Netherlands |  | 1970-2021 | 2017-2019 |
| 16 | Norway |  | 1970-2021 | Not available  (only total number of cause-specific deaths for all ages combined) |
| 17 | Portugal |  | 1970-2021 | 2017-2019 |
| 18 | Spain * |  | 1970-2021 | 2017-2019 |
| 19 | Sweden * |  | 1970-2021 | 2017-2019 |
| 20 | Switzerland * |  | 1970-2021 | 2017-2019 |

Note: ^a^ The WHO MDB did not include death counts for New Zealand for any of the years in the period 2017-19. Instead, we used data from the Health New Zealand Mortality Collection^3^; ^b^ For the period 1970-1989, we treated Germany as two populations, East and West Germany, as Germany was divided into two parts from 1949 to 1990; * denotes the five best performing countries, which form an alternative comparison group for additional (sensitivity) analysis.

### Table S2. List of causes of death and corresponding International Classification of Diseases codes (ICD-10)

| **Cause of death** | **ICD-10 codes** |
| --- | --- |
| Infectious diseases | A00-B99 |
| Neoplasms, incl. | C00-D48 |
| - Lung cancer * | C33-C34 |
| - Other cancers * | rest of C00-D48 |
| Diabetes | E10-E14 |
| Dementia (incl. Alzheimer disease) * | F00-F03, G30 |
| Cardiovascular diseases (CVD), incl. | I00-I99 |
| - IHD * | I20-I25 |
| - Stroke | I60-I64 |
| - Other CVD * | rest of I00-I99 |
| Respiratory diseases * | J00-J99 |
| Substance-related deaths, incl. * | F10–F19, K70-K74, X40–X45, Y10–Y15 |
| - Substance-related disorders | F10–F19 |
| - Poisonings | X40–X45, Y10–Y15 |
| - Alcohol-related liver disease | K70-K74 |
| Injuries * | V01-Y98 (excl. X40–X45, Y10–Y15) |
| - Suicides | X60-X84 |
| - Other injuries | rest of V01-Y98 (excl. X40–X45, Y10–Y15) |
| Other causes of death * | rest |

Note: The sign '*' indicates the categories of causes of death used to construct the main Figure 4. Ill-defined and unknown causes of death (R00-R99) were redistributed proportionally by age, sex, cause of death and country.

### Table S3. Age- and cause-specific contributions (in years) to the differences in life expectancy at birth between each of the English-speaking countries and the average for other HICs, by sex, average for 2017-19.

|  | **FEMALES** | | | | | | **MALES** | | | | | |
| --- | --- | --- | --- | --- | --- | --- | --- | --- | --- | --- | --- | --- |
|  | **Australia** | **Canada** | **Ireland** | **New Zealand** | **UK** | **USA** | **Australia** | **Canada** | **Ireland** | **New Zealand** | **UK** | **USA** |
|  | **All ages** | | | | | | | | | | | |
| **ALL CAUSES** | **0.54** | **-0.42** | **-0.49** | **-1.04** | **-1.52** | **-3.24** | **1.16** | **0.05** | **0.44** | **0.14** | **-0.53** | **-3.56** |
| Infectious diseases | 0.03 | 0.01 | 0.07 | 0.07 | 0.04 | -0.15 | 0.05 | 0.03 | 0.10 | 0.09 | 0.06 | -0.16 |
| Lung cancer | 0.01 | -0.27 | -0.13 | -0.15 | -0.18 | -0.14 | 0.21 | 0.04 | 0.09 | 0.23 | 0.08 | 0.08 |
| Other cancers | 0.18 | 0.14 | -0.23 | -0.13 | -0.24 | 0.16 | 0.21 | 0.32 | 0.05 | 0.09 | -0.02 | 0.34 |
| Diabetes | -0.04 | -0.01 | 0.06 | -0.08 | 0.06 | -0.16 | -0.05 | -0.05 | 0.05 | -0.09 | 0.08 | -0.21 |
| Dementia | 0.00 | -0.16 | -0.04 | -0.11 | -0.47 | -0.26 | 0.01 | -0.10 | -0.01 | -0.05 | -0.23 | -0.09 |
| Respiratory diseases | -0.07 | -0.11 | -0.47 | -0.19 | -0.61 | -0.48 | 0.12 | 0.07 | -0.18 | 0.11 | -0.39 | -0.25 |
| IHD | -0.05 | -0.15 | -0.24 | -0.34 | -0.14 | -0.50 | -0.01 | -0.20 | -0.45 | -0.47 | -0.35 | -0.70 |
| Stroke | 0.01 | 0.07 | 0.00 | -0.12 | -0.05 | -0.01 | 0.09 | 0.10 | 0.06 | 0.05 | 0.01 | 0.02 |
| Other CVD | 0.52 | 0.53 | 0.35 | 0.22 | 0.35 | -0.28 | 0.55 | 0.44 | 0.35 | 0.28 | 0.35 | -0.34 |
| Substance-related disorders | 0.02 | -0.01 | 0.03 | 0.02 | 0.01 | -0.03 | 0.07 | 0.02 | 0.09 | 0.09 | 0.05 | -0.03 |
| Alcohol-related liver disease | 0.03 | -0.02 | 0.02 | 0.06 | -0.07 | -0.10 | 0.09 | 0.04 | 0.08 | 0.18 | -0.04 | -0.08 |
| Poisonings | -0.07 | -0.18 | -0.06 | -0.01 | -0.10 | -0.35 | -0.14 | -0.43 | -0.11 | -0.01 | -0.21 | -0.68 |
| Suicides | -0.04 | -0.03 | 0.04 | -0.06 | 0.05 | -0.03 | -0.14 | -0.09 | 0.05 | -0.14 | 0.08 | -0.20 |
| Other injuries | 0.01 | -0.06 | 0.16 | -0.09 | 0.05 | -0.29 | 0.06 | -0.04 | 0.27 | -0.17 | 0.14 | -0.69 |
| Other causes of death | -0.02 | -0.17 | -0.04 | -0.14 | -0.22 | -0.63 | 0.05 | -0.11 | 0.01 | -0.05 | -0.13 | -0.55 |
|  | **Between ages 0 and 50** | | | | | | | | | | | |
| **ALL CAUSES** | **-0.15** | **-0.46** | **-0.03** | **-0.43** | **-0.35** | **-1.23** | **-0.29** | **-0.68** | **0.05** | **-0.51** | **-0.43** | **-2.06** |
| Infectious diseases | 0.00 | -0.01 | 0.01 | 0.01 | -0.01 | -0.04 | 0.01 | 0.00 | 0.01 | 0.01 | 0.00 | -0.05 |
| Lung cancer | 0.01 | 0.01 | 0.00 | 0.00 | 0.00 | 0.00 | 0.01 | 0.01 | 0.01 | 0.01 | 0.01 | 0.01 |
| Other cancers | 0.02 | 0.01 | -0.01 | -0.05 | -0.04 | -0.02 | 0.01 | 0.01 | 0.01 | 0.00 | -0.01 | -0.01 |
| Diabetes | -0.01 | -0.01 | 0.00 | -0.01 | -0.01 | -0.04 | -0.01 | -0.02 | 0.01 | -0.02 | -0.01 | -0.05 |
| Dementia | 0.00 | 0.00 | 0.00 | 0.00 | 0.00 | 0.00 | 0.00 | 0.00 | 0.00 | 0.00 | 0.00 | 0.00 |
| Respiratory diseases | -0.01 | -0.01 | -0.01 | -0.01 | -0.04 | -0.06 | -0.01 | 0.00 | 0.01 | -0.01 | -0.04 | -0.05 |
| IHD | -0.01 | -0.01 | 0.00 | -0.01 | -0.02 | -0.04 | -0.03 | -0.02 | -0.03 | -0.05 | -0.05 | -0.09 |
| Stroke | 0.00 | 0.00 | 0.00 | -0.01 | -0.01 | -0.01 | 0.00 | 0.00 | 0.01 | 0.00 | 0.00 | -0.01 |
| Other CVD | 0.00 | -0.01 | 0.00 | -0.01 | -0.02 | -0.10 | 0.01 | 0.01 | 0.01 | -0.02 | -0.02 | -0.14 |
| Substance-related disorders | 0.00 | -0.01 | 0.01 | 0.01 | 0.00 | -0.02 | 0.02 | 0.01 | 0.03 | 0.03 | 0.01 | -0.02 |
| Alcohol-related liver disease | 0.00 | -0.01 | -0.01 | 0.01 | -0.05 | -0.04 | 0.01 | 0.00 | -0.01 | 0.03 | -0.05 | -0.05 |
| Poisonings | -0.05 | -0.15 | -0.03 | -0.01 | -0.08 | -0.27 | -0.12 | -0.35 | -0.10 | 0.00 | -0.19 | -0.55 |
| Suicides | -0.05 | -0.05 | 0.01 | -0.08 | 0.02 | -0.04 | -0.16 | -0.11 | 0.00 | -0.17 | 0.02 | -0.18 |
| Other injuries | -0.02 | -0.06 | 0.04 | -0.09 | 0.00 | -0.26 | -0.02 | -0.09 | 0.12 | -0.21 | 0.03 | -0.61 |
| Other causes of death | -0.02 | -0.15 | -0.04 | -0.16 | -0.11 | -0.28 | -0.02 | -0.14 | -0.02 | -0.11 | -0.12 | -0.27 |
|  | **Above age 50** | | | | | | | | | | | |
| **ALL CAUSES** | **0.69** | **0.03** | **-0.46** | **-0.61** | **-1.17** | **-2.01** | **1.45** | **0.73** | **0.39** | **0.64** | **-0.09** | **-1.49** |
| Infectious diseases | 0.03 | 0.02 | 0.06 | 0.06 | 0.04 | -0.11 | 0.04 | 0.03 | 0.08 | 0.08 | 0.06 | -0.11 |
| Lung cancer | 0.01 | -0.28 | -0.14 | -0.15 | -0.18 | -0.14 | 0.20 | 0.03 | 0.08 | 0.22 | 0.08 | 0.07 |
| Other cancers | 0.16 | 0.12 | -0.22 | -0.08 | -0.20 | 0.18 | 0.20 | 0.31 | 0.04 | 0.09 | -0.01 | 0.35 |
| Diabetes | -0.03 | 0.00 | 0.06 | -0.06 | 0.07 | -0.12 | -0.04 | -0.03 | 0.04 | -0.06 | 0.09 | -0.16 |
| Dementia | 0.00 | -0.16 | -0.04 | -0.11 | -0.47 | -0.26 | 0.01 | -0.10 | -0.01 | -0.05 | -0.23 | -0.09 |
| Respiratory diseases | -0.05 | -0.09 | -0.46 | -0.18 | -0.57 | -0.42 | 0.13 | 0.07 | -0.19 | 0.11 | -0.35 | -0.20 |
| IHD | -0.04 | -0.15 | -0.24 | -0.33 | -0.13 | -0.46 | 0.02 | -0.18 | -0.42 | -0.43 | -0.30 | -0.61 |
| Stroke | 0.01 | 0.07 | 0.00 | -0.10 | -0.04 | 0.00 | 0.09 | 0.09 | 0.06 | 0.06 | 0.01 | 0.03 |
| Other CVD | 0.53 | 0.54 | 0.34 | 0.24 | 0.38 | -0.17 | 0.53 | 0.43 | 0.34 | 0.30 | 0.37 | -0.21 |
| Substance-related disorders | 0.02 | 0.00 | 0.02 | 0.02 | 0.01 | -0.01 | 0.05 | 0.02 | 0.06 | 0.06 | 0.04 | -0.01 |
| Alcohol-related liver disease | 0.04 | -0.01 | 0.03 | 0.05 | -0.03 | -0.05 | 0.08 | 0.04 | 0.09 | 0.15 | 0.01 | -0.04 |
| Poisonings | -0.02 | -0.03 | -0.02 | 0.00 | -0.02 | -0.08 | -0.02 | -0.08 | -0.01 | -0.01 | -0.03 | -0.14 |
| Suicides | 0.01 | 0.01 | 0.03 | 0.01 | 0.03 | 0.01 | 0.02 | 0.02 | 0.05 | 0.03 | 0.06 | -0.02 |
| Other injuries | 0.03 | 0.00 | 0.12 | 0.00 | 0.05 | -0.03 | 0.08 | 0.04 | 0.14 | 0.04 | 0.11 | -0.08 |
| Other causes of death | 0.00 | -0.02 | 0.00 | 0.03 | -0.11 | -0.35 | 0.07 | 0.02 | 0.03 | 0.06 | -0.01 | -0.28 |

Note: negative contributions are shown in red colour

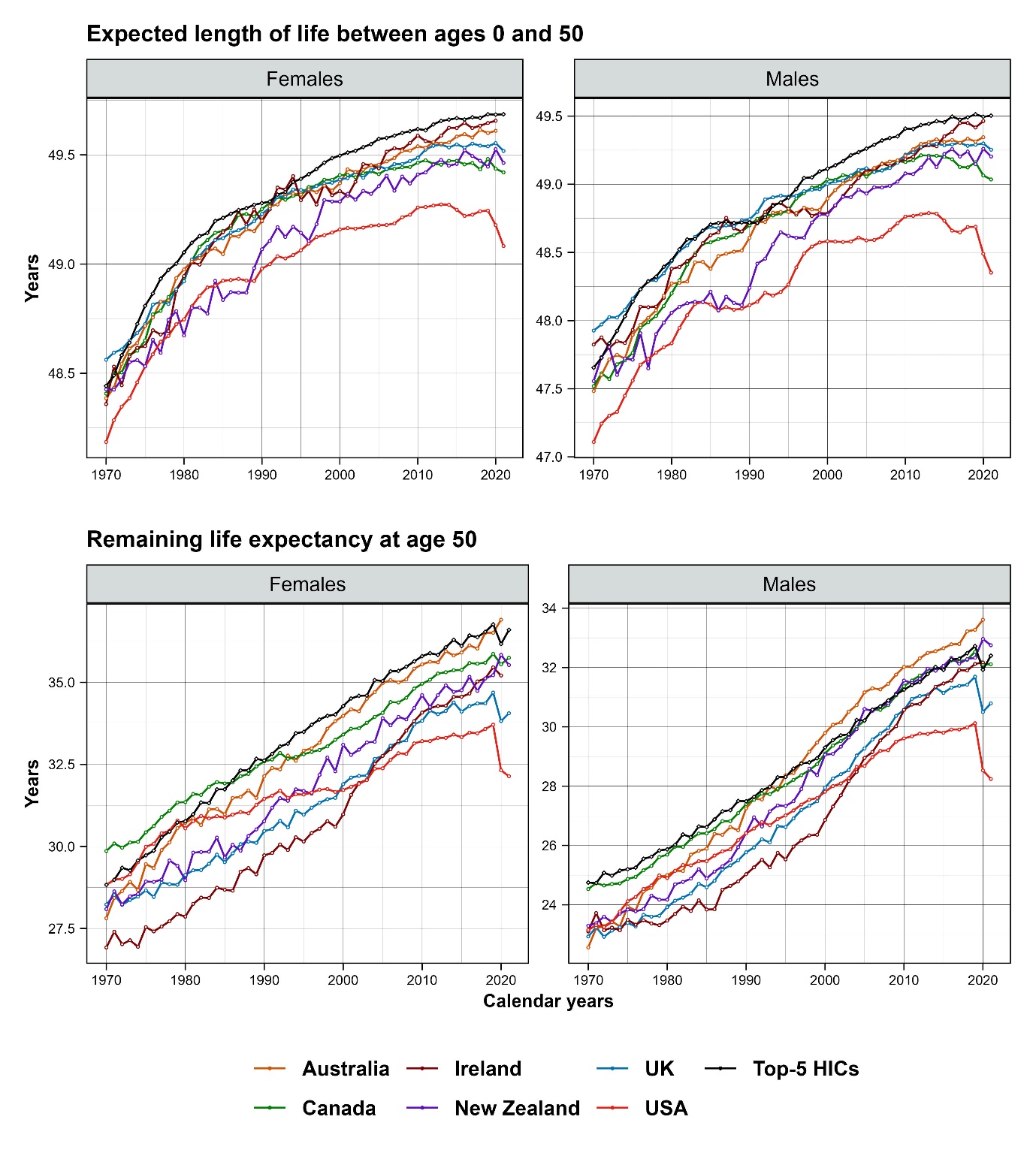

### Figure S1. Time trends in partial life expectancies in English-speaking countries and the average for five best performing HICs, by sex, 1970-2021.

Note: ^a^ life expectancy at birth is the sum of life expectancies between ages 0 and 50 (upper panel), and remaining life expectancy at age 50 (lower panel) multiplied by the probability of surviving up to age 50 (see Supplementary Methods); ^b^ HMD, used as the primary data source for the calculations, does not contain the data for 2021 for Australia and Ireland

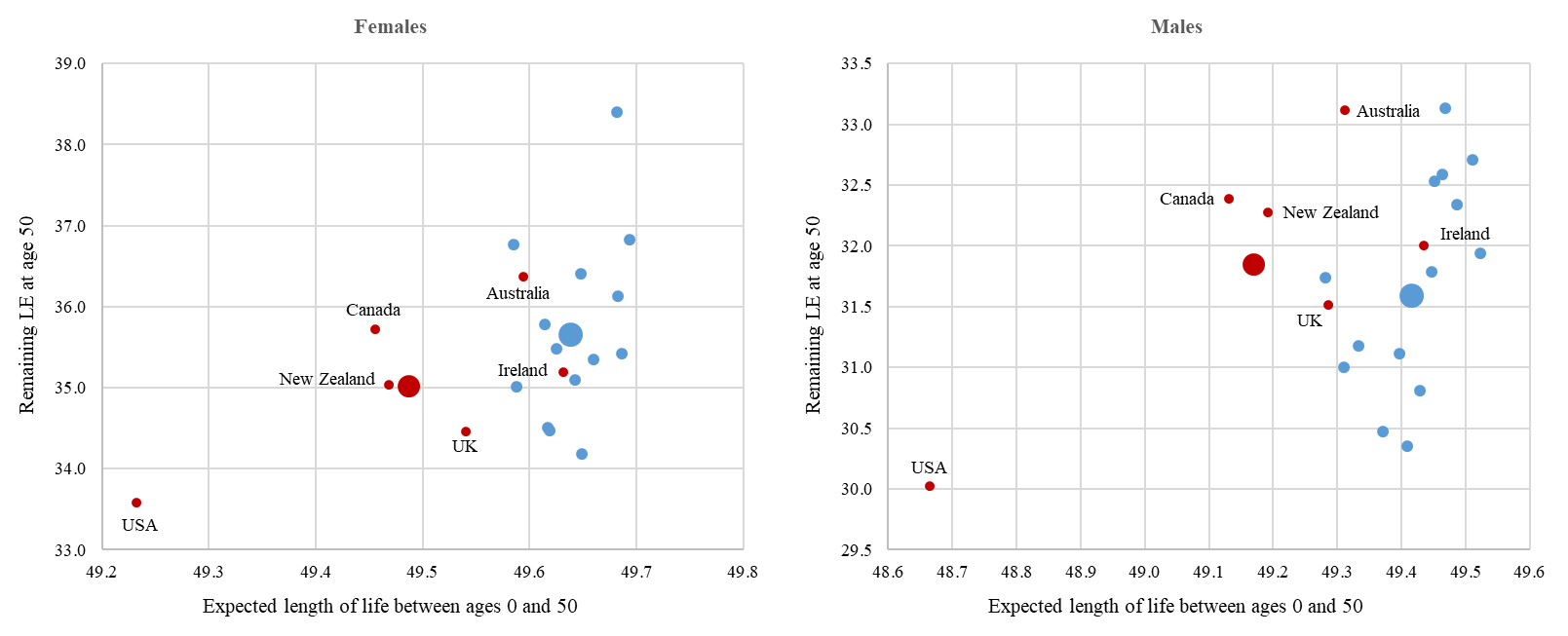

### Figure S2. Association between life expectancy between ages 0 and 50 years and remaining life expectancy at age 50 years, across the English-speaking and 14 other HICs, by sex, average for 2017-19.

Note. Bigger bubbles correspond to the average of the groups (English-speaking countries (in red) and other HICs (in blue)).

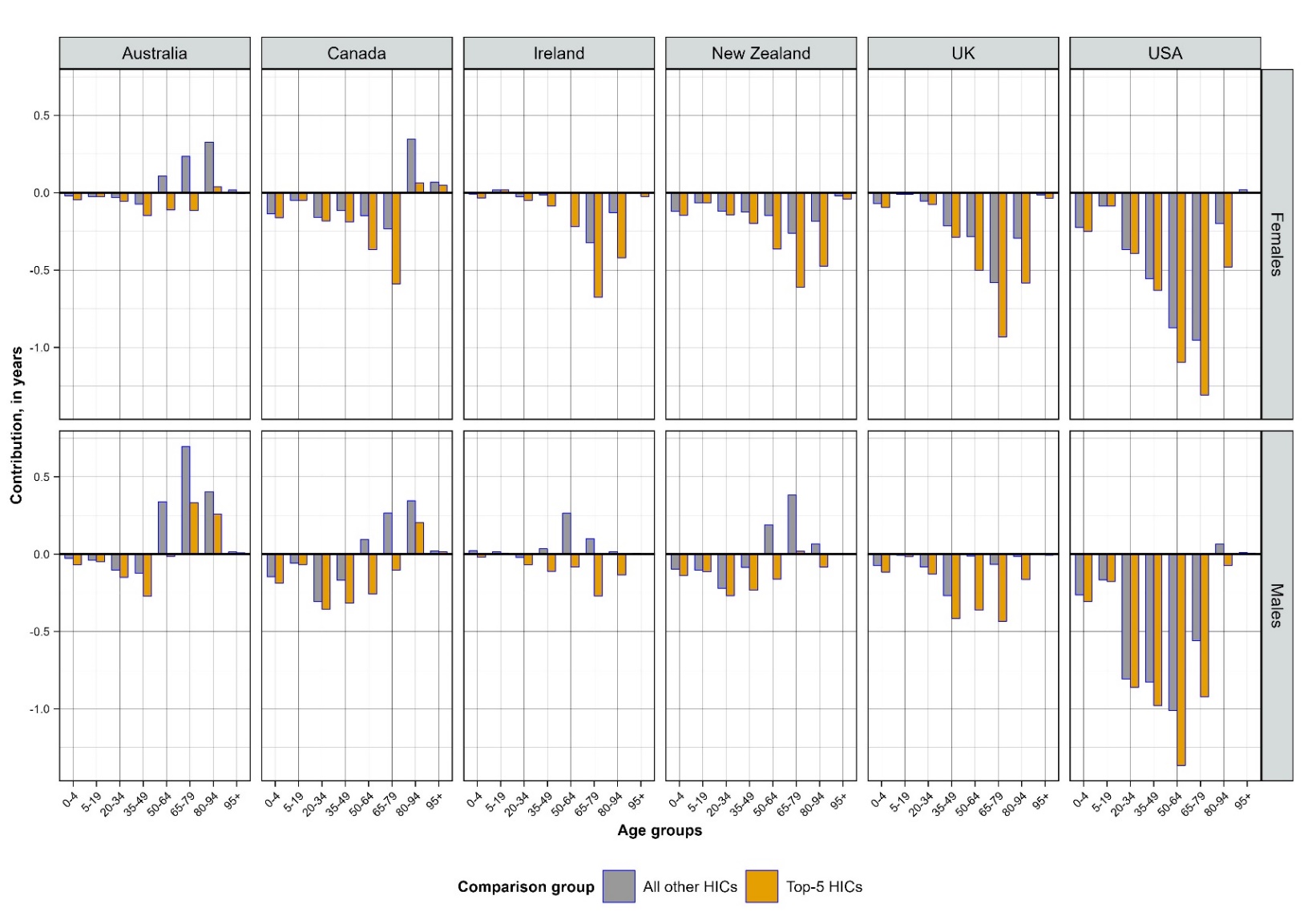

### Figure S3. Age-specific contributions to the gap in life expectancy at birth between English-speaking countries and two comparator groups (average of 14 other HICs and average of 5 best performing other HICs), by sex, in years.

### Supplementary references

1.	HMD. Human Mortality Database. Max Planck Institute for Demographic Research (Germany), University of California, Berkeley (USA), and French Institute for Demographic Studies (France). [Internet]. 2023. Available from: www.mortality.org.

2.	WHO. WHO Mortality Database [Internet]. 2023. Available from: https://www.who.int/data/data-collection-tools/who-mortality-database

3.	Health New Zealand. The Health New Zealand Mortality Collection [Internet]. 2023. Available from: https://tewhatuora.shinyapps.io/mortality-web-tool/

4.	Pascariu M, Rizzi S, Schoeley J, Danko M. Package ‘ ungroup ’ [version 1.4.2]. 2022; Available from: https://cran.r-project.org/web/packages/ungroup/ungroup.pdf

5.	Wilmoth J, Andreev K, Jdanov J, Glei D, Riffe T. Methods Protocol for the Human Mortality Database (Version 6). *Available at www.mortality.org*. 2021;
